# A 5’ UTR CCG expansion in *TBC1D7* causes oculopharyngodistal myopathy

**DOI:** 10.64898/2026.03.27.26349107

**Authors:** Liedewei Van de Vondel, Riccardo Curro, Stefano Facchini, Isaac R. L. Xu, Jonathan De Winter, Ilaria Quartesan, Alice Monticelli, Alicia Alonso-Jimenez, Willem De Ridder, Alessandro Bertini, Gustavo Alves, Francesca Pizzuto, Hermione Ugolini, David Pellerin, Tim De Pooter, Ashirwad Merve, Pedro Machado, OPDM study group, Lydia Sagath, Kornelia Neveling, Alexander Hoischen, Michael G Hanna, Robert D. S. Pitceathly, Henry Houlden, Arianna Tucci, Enrico Bugiardini, Stefen Brady, Mark Roberts, Matt C. Danzi, Stephan Züchner, Jonathan Baets, Andrea Cortese

## Abstract

Oculopharyngodistal myopathy (OPDM) is a group of rare, hereditary myopathies characterized by ptosis, external ophthalmoplegia, facial, pharyngeal and distal limb weakness and classically with rimmed vacuoles and intranuclear inclusions on muscle biopsy. Heterozygous CCG-CGG repeat expansions in the 5’ UTR of six genes are known to cause OPDM, only one of which (*ABCD3*) has been reported in individuals of European ancestry. Here, we identify heterozygous CCG expansions in *TBC1D7*, ranging from 87-134 repeats, in three unrelated families of European and mixed African European descent, establishing *TBC1D7* as a new OPDM gene. Using integrated long-read and short-read sequencing technologies and large population datasets, we define the structure of the *TBC1D7* tandem repeat and show that this locus is strikingly variable in the control population – a recently recognized hallmark of pathogenic repeat loci. We furthermore investigate epigenetic regulation and repeat length variability at the repeat locus, demonstrating CCG repeat methylation as plausible mechanism for the observed non-penetrance in one unaffected individual carrying a large repeat expansion, while in affected patients the repeat is unmethylated. Patient-derived fibroblasts show increased *TBC1D7* expression, and p62-positive intranuclear inclusions are observed on muscle biopsy, supporting a dominant toxic gain-of-function mechanism analogous to other CCG-expansion disorders. This study expands the known genetic architecture of OPDM and distal myopathies in general and reinforces the emerging paradigm in which the sequence motif and genomic context of repeat expansions, rather than gene function alone, are key drivers of disease. The identification of *TBC1D7* as a repeat-expansion myopathy gene further highlights the need for systematic interrogation of noncoding repeat loci in unresolved neuromuscular disease cohorts.

## Introduction

Oculopharyngodistal myopathy (OPDM) is a group of rare hereditary muscle diseases characterized by progressive symptoms, including adult-onset ptosis, external ophthalmoplegia, and weakness of the facial, pharyngeal and distal limb muscles. A hallmark feature of OPDM is the presence of rimmed vacuoles in the muscle biopsy^1,2^. Although long suspected to be of genetic origin, only recently heterozygous CCG-CGG repeat expansions in the 5’ UTR of six genes have been identified as a cause of OPDM^3–8^. While OPDM1-4 (*LRP12, GIPC1, NOTCH2NLC and RILPL1,* respectively) and OPDM6 (*LOC642361/NUTM2B-*AS1) were all identified in patients of Asian descent, OPDM5 (*ABCD3*) was identified in families of European ancestry, representing the first genetic cause of OPDM in non-Asian populations^7^. Despite these advances and identified genetic loci, many unsolved OPDM patients remain, indicating additional genetic heterogeneity. The consistent association between CCG-CGG repeat motifs and the OPDM phenotype further supports the rationale for continued searches for novel repeat expansions with this motif^9^.

In this study, we sought to uncover the genetic basis of OPDM in multiple families of European ancestry using both short-read and long-read whole genome sequencing. We studied three unrelated families comprising seven affected individuals and identified CCG repeat expansions in the 5′ UTR of *TBC1D7* as the shared genetic cause of OPDM.

## Materials & Methods

### Patient Cohort and Clinical Studies

All experiments and clinical studies were approved by the respective ethical committees of the University of Antwerp, the Antwerp University Hospital and the University College London (Northeast-Newcastle & North Tyneside Research Ethics Committee, 22/NE/0080, Ethics Committee UZA/UAntwerp, B300201525715). All individuals provided informed consent. Patients diagnosed clinically with OPDM, but without a confirmed genetic cause, were recruited at both Antwerp and London centers, and standardized clinical information was collected, including demographics, family history, age at disease onset, initial symptoms, presence of ptosis, ophtalmoparesis, facial weakness, dysphagia, dysarthria, muscle weakness pattern, age at death. Muscle biopsies from the gastrocnemius were performed for individuals A:II:1 and C:II:1, followed by routine histopathology including H&E, Gomori trichome, NADH and p62 staining. Ultrastructural studies were available for the muscle biopsy of A:II:1. Electromyography (EMG) was performed for individuals A:I:1, A:II:1, and C:II:1. Muscle magnetic resonance imaging (MRI) was performed for patients A:I:1 and A:II:1, including a longitudinal follow-up in individual A:II:1.

### Short read (sr)-WGS analysis in the Genomics England 100,000 genome project

The 100,000 Genomes Project (100K GP), run by Genomics England, was established to sequence whole genomes of patients of the National Health Service (NHS) of the UK, affected by rare diseases and cancer^10^. We analysed a cohort of 371 myopathy cases and 35,376 non-neurological controls enrolled in the 100K GP – Rare Disease Cohort. For all individuals srWGS was available (paired-end, 150 bp, average read depth >30×). All samples were probed with ExpansionHunter denovo (EHdn) v0.9.0 to obtain a count of anchored in-repeat reads, then jointly analysed using the “outlier” method provided in the EHdn package^11^. The method computes a z-score for each sample, based on the original distribution of read counts. The score is to be interpreted as distance to the median, relative to the distribution width.

### Long read (lr)-WGS and Bionano optical genome mapping in the patient cohort

PacBio HiFi sequencing for A:I:1, A:I:2, A:II:1 and A:II:2 and Bionano optical genome mapping (OGM) on blood-derived DNA for A:II:1 was performed as part of a larger study in the Solve-RD consortium^12^. Genome-wide structural variant calling using pbsv v2.4.1 was performed for family A as described. OGM was generated as previously described^13^. Following identification of the *TBC1D7* locus, Oxford Nanopore Technologies (ONT) PromethION sequencing data was generated for DNA extracted from muscle biopsy of A:II:1, and from DNA extracted from peripheral blood for A:II:1, B:III:2, C:I:1 and C:II:1, as well as OGM for B:III:2. For muscle-biopsy derived DNA from A:II:1, DNA was extracted from 100 mg of muscle biopsy using the QIAamp DNA mini kit (Cat° 51304) and transferred to the Genomics Core Leuven for ONT Promethion sequencing. For DNA from peripheral blood for B:III:2, C:I:1 and C:II:1, ONT PromethION sequencing was performed at the Long Read Facility of UCL Institute of Neurology. ONT PromethION reads were basecalled using Dorado (Oxford Nanopore Technologies, version 1.3.1), enabling analysis and visualization of methylation status. From generated data, the *TBC1D7* tandem repeat (TR) locus (chr6:13328476-13328603, GRCh38) was called using STRdust^14^. Sequence composition analysis and visualization was performed using the RepeatXuTRACTR algorithm^15^.

### Repeat primed (RP)-PCR

RP-PCR was performed to study segregation of CCG expansion in families B and C, as well as to screen expansion of the *TBC1D7* locus in a larger cohort of 27 individuals affected by OPDM from the UK, France, Turkey and Italy. The following primers were used: *TBC1D7* Fw: FAM-CAGACGGCCCGGGAGACAAA; anchor: CAGGAAACAGCTATGACC; *TBC1D7* Rv: CAGGAAACAGCTATGACCCGGCGGCGGCGG. The PCR mix contained 1.25 U Takara LaTaq (Takara, Shiga, Japan), 1× GC Buffer II, 600 μM each dNTP mixture, 0.4 μM of each primer, 1M Betaine (Sigma Aldrich), <1 μg DNA, and ddH2O for a final volume of 25 μL. The following thermal conditions were used: initial denaturation at 95 °C for 5 minutes, followed by 50 cycles of 95 °C for 30 seconds, 98 °C for 10 seconds, 58 °C for 30 seconds, 72 °C for 2 minutes; final extension of 72 °C for 5 minutes. Fragment analysis was conducted on an ABI-3730XL Sequencer, and data were visualised using Geneious Prime (Biomatters Ltd).

### Analysis of the *TBC1D7* locus in the control population

The *TBC1D7* TR locus (chr6:13328476-13328603, GRCh38) was genotyped in 1,020 publicly available ONT datasets from the 1000 Genomes consortium^16^ using STRdust^14^. 1000 Genomes was accessed on 24th of June 2025 from https://registry.opendata.aws/1000-genomes. TR alleles were decomposed and visualized using the RepeatXuTRACTR algorithm^15^. Additionally, all repeat loci from the Adotto catalogue^17^ that have a CCGCTG composition as their most frequent longest pure segment (LPS)^18^ were genotyped and decomposed in the 1000 genomes ONT dataset, allowing to construct a distribution of LPS length of CCGCTG per loci in the control population.

### Haplotype analysis

Blood-derived PacBio HiFi or ONT data from A:I:1, A:II:1, B:III:2, C:I:1, C:II:1 was haplotyped using Whatshap^19^. Bam files were split into the two called haplotypes per patient. SNPs were called using bcftools mpileup and bcftools call in 100kb around the TR locus. SNPs were consequently filtered to quality ≥ 30, depth ≥ 10, mapping quality ≥ 40, minor allele frequency ≥ 5% based on GnomAD V4, resulting in 107 high-quality, frequent SNPs.

### Fibroblast culture and quantitative PCR for *TBC1D7* expression

Human-derived skin fibroblasts from patients B:III:2, C:II:1, and age-matched controls were cultured in Dulbecco’s Modified Eagle Media (DMEM) supplemented with 10% FBS at 37°C in a 5% CO_2_ humidified incubator. Total RNA was extracted from fibroblasts using the RNeasy Mini kit (Qiagen). cDNA was synthesized from 500 ng of total RNA with Maxima H Minus cDNA Synthesis Master Mix (Invitrogen). Real-Time quantitative PCR was performed using the TaqMan Fast Advanced Master Mix. *TBC1D7* expression level (probe Hs00212898_m1) was normalized to TATA-binding protein (*TBP*) (Hs00427620_m1). Amplified transcripts were quantified using the comparative CT method and presented as normalized fold expression change (2-ddCt).

## Results

### Identification of the 5’ UTR *CCG-*expansion in *TBC1D7*

In the HiFi data of family A, generated within the Solve-RD framework^12^, we conducted a genome-wide search for SVs affecting exons or UTRs that segregated with disease. During a Solve-RD Solvathon^20^, this analysis identified an insertion in the 5′ UTR of *TBC1D7*, that, upon closer inspection of the reads, was found to consist of a CCG TR expansion in both individuals A:I:1 and A:II:1 and represented the only exon– or UTR-affecting SV segregating across all four sequenced family members. The *TBC1D7* TR locus (chr6:13328476-13328603, GRCh38) was recalled using TRGT^21^ to enable size estimation of the repeat, at 84 units for A:I:1 while read depth (only two reads spanning the repeat) was insufficient to allow accurate repeat estimation in A:II:1. Therefore, to obtain a more accurate size estimate in A:II:1, we performed ONT sequencing of fresh peripheral blood DNA, ascertaining a median expansion length of 113 CCG units across 20 reads (Figure 1A). Moreover, optical genome mapping (OGM) in A:II:1 provided further orthogonal confirmation of the presence of an expansion at the *TBC1D7* repeat locus (Supplementary Figure 1A).

**Figure 1:**
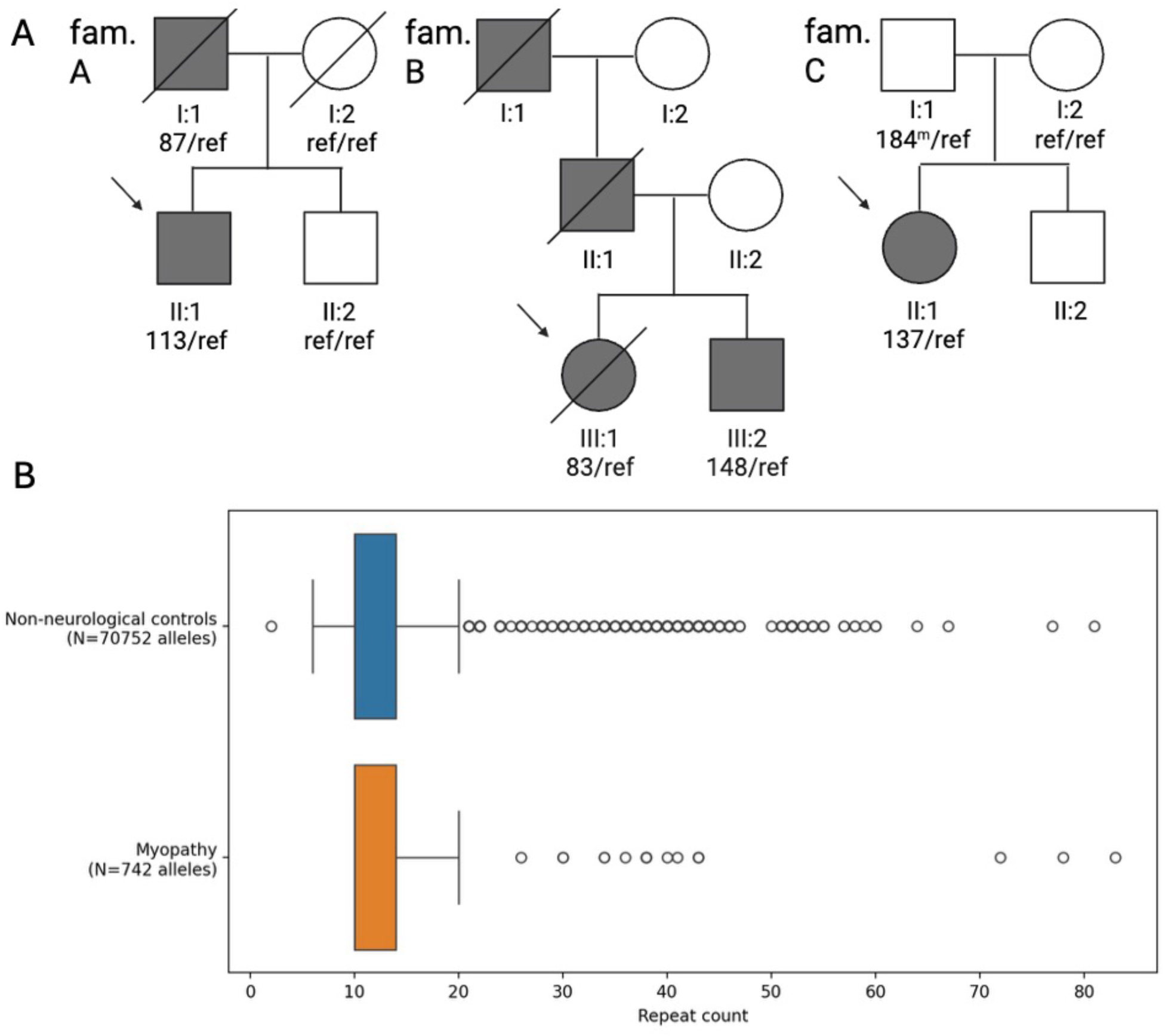
Identification of the TBC1D7 5’ UTR CCG repeat expansion. (A) Pedigrees of three aFected families (fam. A-C) carrying the repeat expansion. AFected individuals are coloured black, repeat sizes in CCG units for expanded alleles are indicated below each individual, as well as the reference (ref) genotype. (B) Identification of the TBC1D7 CCG expansion causing OPDM in SR-WGS through Genomics England 100,000 Genome Project.

Independently, the *TBC1D7* repeat was identified via an unbiased analysis of the Genomics England 100K GP – Rare Disease Cohort srWGS data. In particular, we used the outlier method in EHdn to prioritise loci containing CCG-CGG expansions, which were enriched in 371 myopathy cases compared to 35,376 controls. The method reports a top z-score for each locus, representing the largest expansion in patients relative to the median in controls. We identified three CCG loci showing a high z-score in myopathy cases, including *ABCD3, FRA10AC1,* and *TBC1D7* (z score = 23.5, 14.4 and 14.4, respectively). Notably the gene with the highest z-score, *ABCD3*, was already identified using this method and reported to be disease causing^7^.

We then applied ExpansionHunter v5 to obtain more accurate size estimates for *FRA10AC1 and TBC1D7* expansions in the srWGS data. *FRA10AC1* was excluded because 117 control individuals carried expansions larger than the maximum observed in myopathy cases (97 CCG units, Supplementary Figure 1C), while the large z-score likely reflected the marked variability of this locus in the general population^18^. On the other hand, we confirmed the presence of a large monoallelic CCG expansion in the 5’ UTR of *TBC1D7* in three OPDM cases, B:III:1, B:III:2 and C:II:1 (Figure 1A) from two unrelated families, with initial srWGS based size estimates of 84, 79 and 75 CCG respectively, while large expansions were rare in controls (Figure 1B). Repeat-primed PCR (RP-PCR) was then performed in the unaffected parents of patient C:II:1 showing paternal inheritance of the expanded allele (Supplementary figure 1D). We next generated ONT data for affected probands B:III:2 and C:II:1, and unaffected individual C:I:1, showing median repeat size, at 148, 137 and 184 CCG units respectively (Figure 1A). OGM provided orthogonal confirmation of repeat presence in B:III:2 (Supplementary Figure 1B).

Median pathogenic repeat lengths range from (CCG)_83-148_ across identified patients, with incomplete penetrance of the repeat present in C:I:1 at a median repeat length of (CCG)_184._ (Figure 1A). No repeat expansions at the previously reported OPDM loci (*LRP12, GIPC1, NOTCH2NLC, RILPL1, ABCD3, NUTM2B-AS1*) were identified in any affected individual, nor were there any other candidate TR, single nucleotide or SV candidates that matched segregation. An additional cohort of 27 OPDM index patients from the UK, France, Italy and Turkey was screened by RP-PCR, all of whom tested negative for the *TBC1D7* repeat expansion.

### Single-molecule methylation profiling and repeat instability

Since methylation-dependent modulation of penetrance has previously been described for CCG expansions^2,5,7,22,23^, and individual C:I:1 exhibited non-penetrance despite a large CCG expansion, we next examined methylation at the repeat locus. Interestingly, the expanded repeat and the flanking CpG island in the promoter region of *TBC1D7* in the unaffected individual C:I:1 were fully methylated, a pattern predicted to suppress gene expression. Among 17 reads fully spanning the locus, 13 were hypermethylated (Figure 2). The remaining four reads were unmethylated and corresponded to shorter repeat lengths (<318 bp; 108 repeat units). Hypermethylation was not observed in reads from other affected individuals.

**Figure 2:**
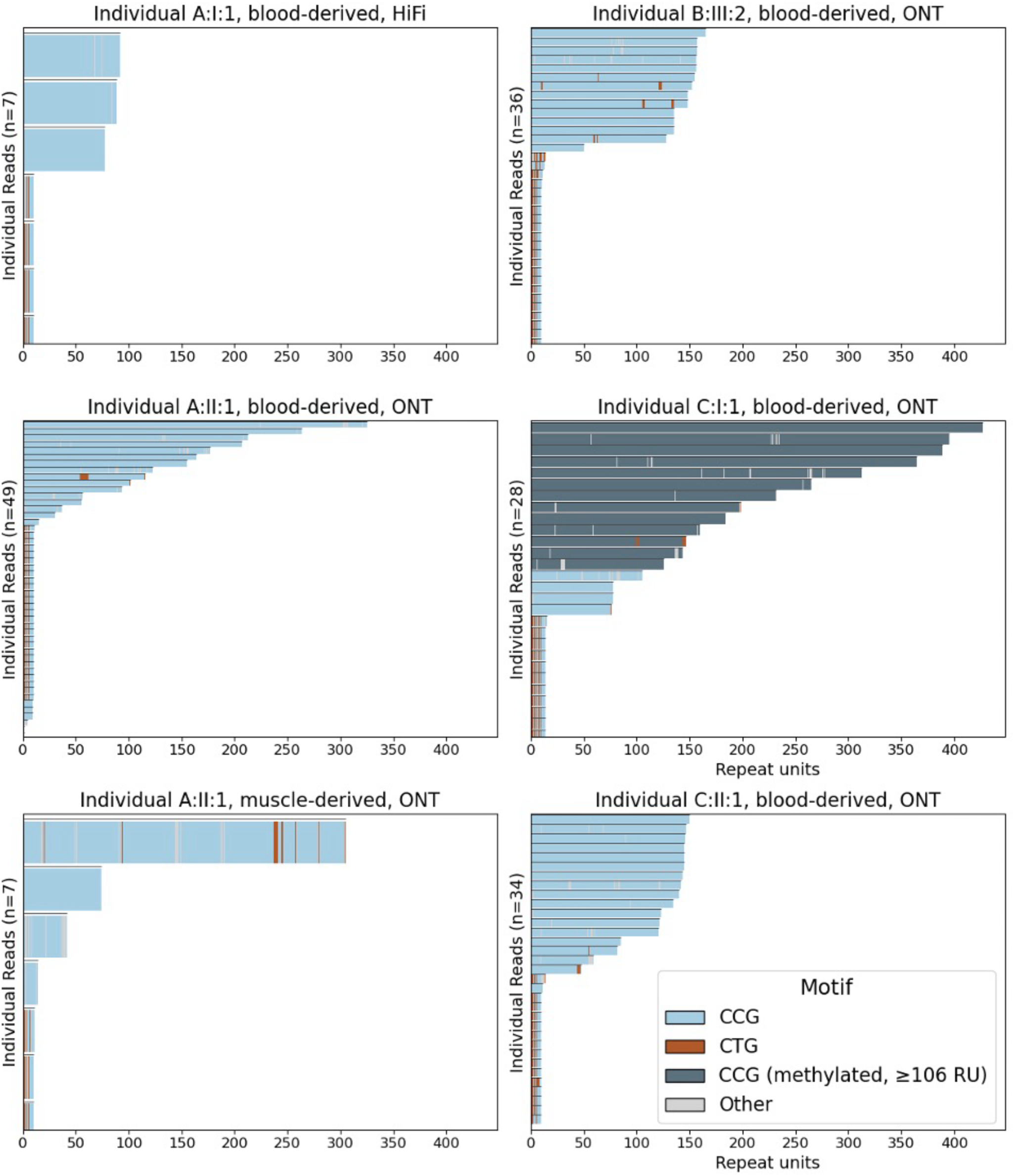
Single-molecule characterization of TBC1D7 repeat lengths across individuals, tissues, and sequencing platforms. Per-read repeat lengths for the expanded TBC1D7 allele in aFected individuals, shown for HiFi and ONT sequencing. Each bar represents a single aligned read, ordered by read length. Light blue indicates unmethylated CCG tracts, while dark blue marks methylated reads.

Affected individuals showed limited repeat instability in blood-derived ONT data, with relatively low inter-read variability observed in A:I:1, B:III:2, and C:II:1 (standard deviations 22, 85, and 98 bp, respectively; Figure 2). In contrast, the unaffected carrier C:I:1, who harboured the largest repeat, showed greater variability (standard deviation 362 bp). Muscle-derived ONT sequencing was additionally performed for A:II:1, but yielded only three spanning reads, resulting in suboptimal coverage that precluded accurate assessment of repeat size, somatic instability, and methylation status. No marked somatic expansion in absolute repeat length was observed compared with blood-derived reads. Inter-read variability was present in both tissues (standard deviation 257 bp in blood and 431 bp in muscle), although these estimates are influenced by the differing number of spanning reads (20 versus three, respectively). Notably, the unaffected parent C:I:1 carried, on average, a larger allele than his affected daughter C:II:1 (median 184 versus 137 CCG units), consistent with paternal contraction described for other CCG repeat loci. In contrast, although the repeat was also transmitted paternally from A:I:1 to A:II:1, it expanded on average during transmission, increasing from a median of 87 CCG units in A:I:1 to 113 CCG units in A:II:1.

### The *TBC1D7* locus shows pathogenic hallmarks in the control population

The *TBC1D7* GRCh38 reference locus has a (CCGCTG)_3_(CCG)_4_ sequence motif, and analysis of the 1000G ONT data revealed that most (1620/1718, 94.3%) alleles in the control population have (CCGCTG) as their longest pure segment (LPS) – that is, the longest uninterrupted stretch of identical repeat units (Figure 3A). The same motif was detected in the non-expanded allele in all patients. Of the 98 alleles with a (CCG)_n_ LPS (median length: (CCG)_9_) the five longest alleles were four individuals carrying (CCG)_12_ and one interrupted (CCG)_40_(CGCACG)(CCG)_18_. None of the pure CCG stretches in control alleles were thus comparable in length to those observed in patients (Supplementary Figure 2). In addition, expanded alleles over 50 repeats were very rare (79/70752, 0.1%) in the 100K GP (Figure 1B).

**Figure 3:**
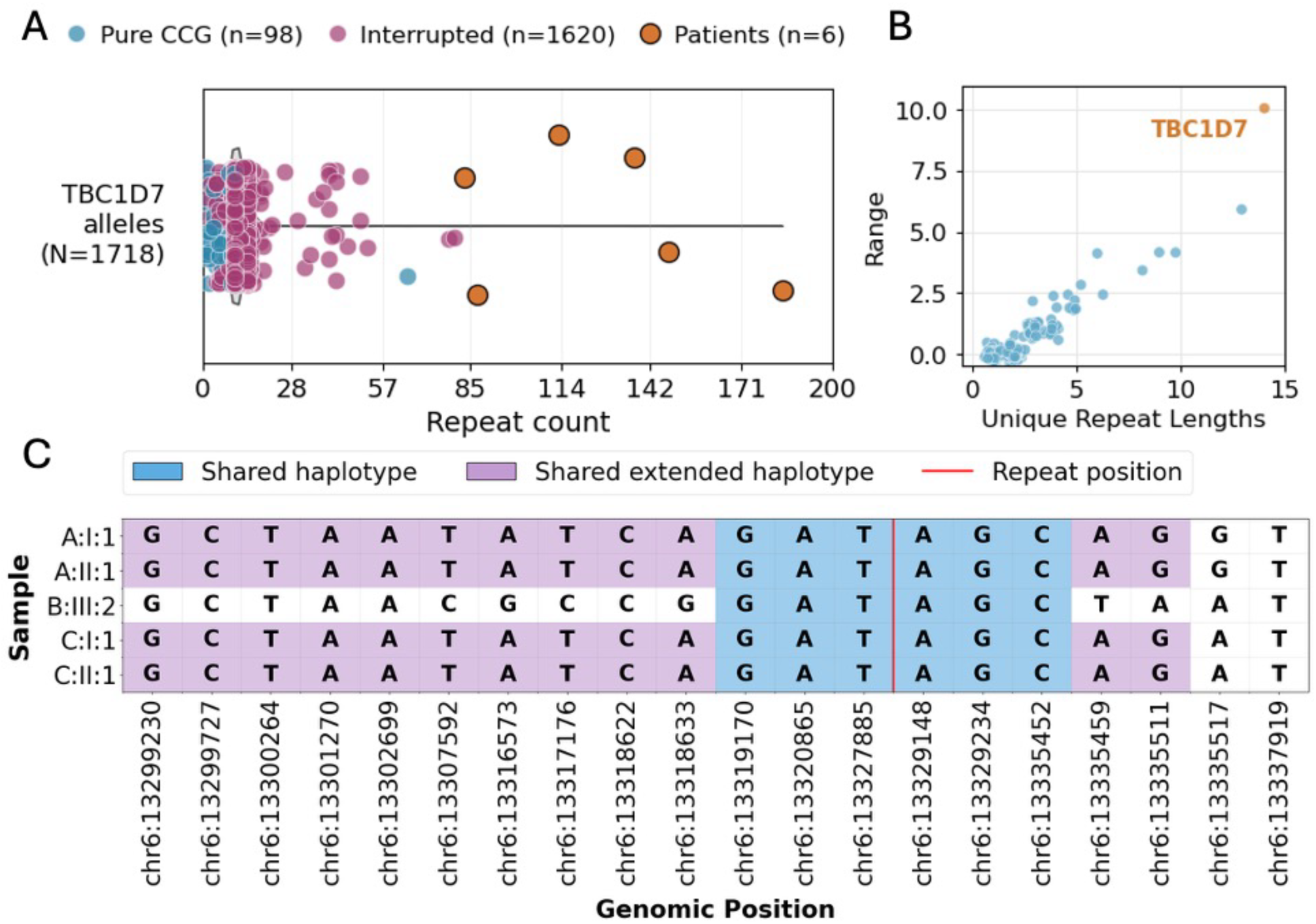
Pathogenic hallmarks of the 5’ UTR CCG expansion in TBC1D7. (A) Comparison of the TBC1D7 locus in 1000G ONT control genomes versus patients. Repeat counts represent total number of trinucleotide units. Alleles that have CCGCTG as LPS are indicated as interrupted in purple, whereas alleles with pure CCG are indicated in blue. Patient CCG repeat counts indicated in red. B) Scatterplot of the number of unique repeat lengths and pure CCGCTG LPS range (99th percentile to median) at all CCGCTG loci in the genome, as characterized in the 1000G ONT genome dataset. TBC1D7 is indicated in red. (C) Phased expanded alleles from aFected individuals in families A–C reveal a shared haplotype spanning the region flanking the TBC1D7 repeat. A core conserved block (blue) encompasses the expansion site (red line), while extended shared segments (purple) are present in families A and C, supporting a common ancestral origin.

Visualization of control alleles from the 1000G revealed substantial inter-individual variability in repeat length, a feature recently proposed as a hallmark of pathogenic tandem repeats^18^. From previous analyses, all known 5’ UTR CCG-CGG loci, including those implicated in OPDM, stand out in terms of repeat length variability when compared to other repeat loci in the genome with the same motif^18^. In contrast to coding variants, where pathogenic loci are typically in genes under evolutionary constraint, disease-associated tandem repeat loci are distinguished by variability, possibly reflecting intrinsic instability and the possibility to expand into pathogenic lengths. Because most *TBC1D7* alleles in the control population have a CCGCTG LPS, this gene was not detected in said analysis. However, when we genotyped all CCGCTG loci in the Adotto catalogue^17^ in the 1000G ONT dataset and quantified the LPS distribution across all alleles, *TBC1D7* emerged as the most variable CCGCTG locus in the genome (Figure 3B).

### Shared haplotype of the *TBC1D7* CCG expansion

Because all previously described OPDM-associated CCG-CGG expansions display a marked ancestry-specificity, and family B was of mixed African European descent while families A and C were of European descent, we wondered whether the *TBC1D7* expansion occurred on a common ancestral haplotype (Figure 3C). Using common variants, with a minor allele frequency of ≥ 0.05 in gnomAD V4, we identified a ∼16kb region (chr6:13,319,170-13,335,452), in which all affected individuals share the same repeat-containing haplotype. In families A and C, this shared segment extends to ∼68kb (chr6:13,267,449-13,335,511), consistent with their closer genealogical relatedness.

### Clinical and histopathological features of individuals carrying the *TBC1D7* CCG expansion

In total, clinical data of five affected individuals carrying *TBC1D7* CCG expansions were available for review (Table 1). While age at onset for B:III:1, B:III:2 and C:II:1 was in the second decade of life, patients from family A became symptomatic in their fifties and forties. Initial symptoms were also diverging, with distal weakness developing before facial symptoms for family A, while ptosis was the presenting symptom for families B and C. At last follow-up, most patients showed a typical OPDM phenotype, with ptosis, facial weakness, dysphagia, and distal weakness, the latter more pronounced in the lower than the upper limbs. Proximal muscles were mildly affected in all cases but one. Patients from family B and C required walking aids since their thirties. The disease was particularly severe in patients B:III:1 and B:III:2, both requiring percutaneous gastrostomy for feeding (sixth and fifth decade, respectively) and non-invasive ventilation due to respiratory involvement (fourth to fifth decade). At last examination (in the sixth decade), patient B:III:2 could communicate only with the help of a voice synthesizer.

**Table 1:** Clinical characteristics of individuals carrying the *TBC1D7* CCG repeat expansion across families A–C. The table summarizes repeat length, sex, age at onset and last follow-up, presenting symptom, disease progression, pattern of muscle involvement (ocular, facial, upper and lower limb distribution), bulbar involvement, muscle biopsy findings (vacuoles and p62-positive inclusions), serum creatine kinase (CK) levels, electromyography (EMG) results, and disability status. Severity annotations (mild, severe) are indicated where available. Abbreviations: UL, upper limb; LL, lower limb; CK, creatine kinase; EMG, electromyography; NIV, non-invasive ventilation; PEG, percutaneous endoscopic gastrostomy.

| Family ID | Repeat Length | Sex | Age at last-follow up (decade) | Age at onset (decade) | First symptom | Progression | Ptosis | Ophthalmoparesis | Facial Weakness | Proximal UL Weakness | Distal UL Weakness | Proximal LL Weakness | Distal LL Weakness | Dysphagia | Dysarthria | Muscle biopsy: vacuoles | Muscle biopsy: p62 inclusions | CK levels (IU/L) | EMG | Disability |
| --- | --- | --- | --- | --- | --- | --- | --- | --- | --- | --- | --- | --- | --- | --- | --- | --- | --- | --- | --- | --- |
| A:I:1 | 87 | Male | Eighth | Sixth | Bilateral foot drop | Slow to moderate | Yes | No | Yes | Yes | No | Yes | Yes(severe) | Yes | No |  |  | 315 | Myopathic |  |
| A:II:1 | 113 | Male | Sixth | Fifth | Bilateral foot drop | Moderate | No | No | No | No | Yes (mild) | Yes | Yes(severe) | Yes | No | Yes | Yes | 852-1288 | Myopathic |  |
| B:III:1 | 83 | Female | Sixth | Second | Ptosis | Slow | Yes | Yes | Yes (severe) | Yes | Yes (severe) | Yes (severe) | Yes (severe) | Yes | Yes | Yes |  | 572 |  | NIV (39), wheelchair (40), PEG (52) |
| B:III:2 | 148 | Male | Sixth | Second | Ptosis | Slow | Yes | Yes (severe) | Yes (severe) | Yes (severe) | Yes (severe) | Yes (severe) | Yes (severe) | Yes | Yes |  |  |  |  | wheelchair (30), PEG (46), NIV (47), bedridden |
| C:II:1 | 137 | Female | Fifth | Second | Ptosis | Slow | Yes | Yes | Yes | Yes | Yes (severe) | Yes (severe) | Yes (severe) | No | Yes (mild) | No (severe atrophy) | Yes | 288 | Myopathic | stick (35), mobility scooter (44) |

Needle EMG showed myopathic changes in all cases investigated and serum creatine kinase levels were mildly to moderately elevated (range: 288-1288 U/L). Muscle MRI was performed in patient A:II:1 and confirmed fatty degeneration of the distal muscles, with predominant involvement of the posterior compartment (Figure 4A). A longitudinal MRI assessment after 15 years demonstrated a further progression of the fatty transformation process, most notably in the posterior compartment of the calf, with additional involvement of the anterior lower-leg muscles (Figure 4B). Muscle biopsy was available for patients A:II:1 and C:II:1, displaying the hallmark rimmed vacuoles (Figure 4C-H). Within the limits of interpretation due to severe atrophy and extensive fibroadipose replacement of the muscle tissue, rare p62-positive intranuclear inclusions were observed in both patients.

**Figure 4:**
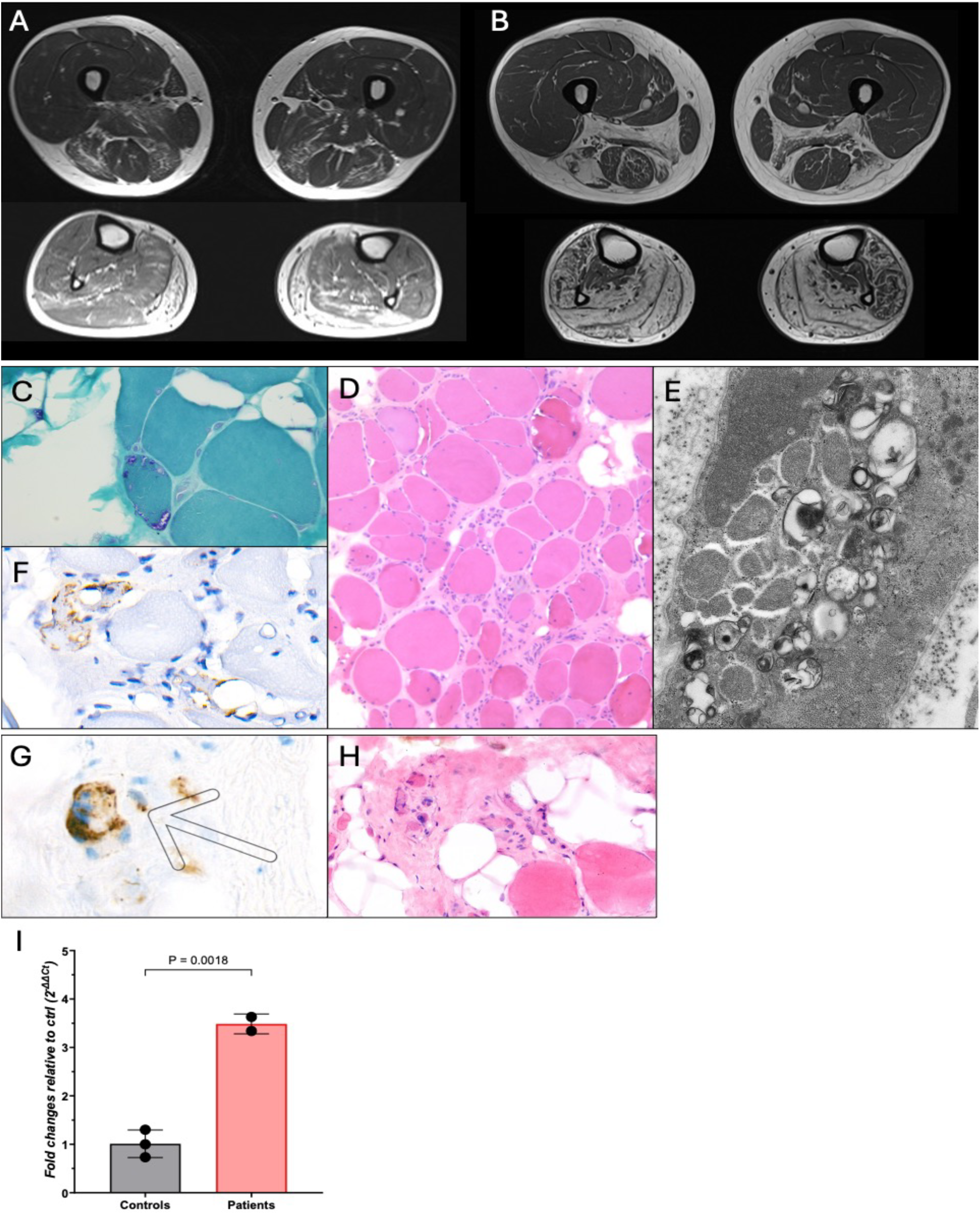
Radiological and pathological features of patients with TBC1D7-related OPDM. (A-B) Muscle MRI at the thigh (upper panels) and calf level (lower panels) of patient A:II:1 in the fifth (A) and sixth decade (B), illustrating longitudinal progression of fatty infiltration, with predominant involvement of the biceps femoris, adductor longus, and posterior calf muscles. (C–F) Muscle biopsy of A:II:1. (C) Gomori trichrome staining showing a subsarcolemmal rimmed vacuoles in an atrophic fiber. (D) Hematoxylin–eosin staining showing marked endomysial fibrosis and fiber size variation. (E) Electron microscopy demonstrating sarcoplasmic tubulofilamentous inclusions. (F) Immunohistochemistry showing p62 positivity at rimmed vacuoles. (G–H) Muscle biopsy of C:II:1. (G) p62 immunohistochemistry showing intranuclear inclusions in addition to sarcoplasmic aggregates. (H) Hematoxylin–eosin staining showing a region of muscle with end-stage fibrosis, fatty replacement, and the presence of small, severely atrophic fibers. (I) TBC1D7 expression in patient-derived fibroblasts. qPCR analysis demonstrates increased TBC1D7 transcript levels in patient fibroblasts compared with controls.

Skin fibroblasts were obtained from patients B:III:2 and C:II:1 and RNA was extracted for qPCR analysis. Notably, *TBC1D7* transcript was significantly overexpressed in patients compared to three age– and sex-matched healthy controls (Figure 4I).

## Discussion

Here, we describe the identification of a CCG expansion in the 5′ UTR of *TBC1D7* as a novel cause of OPDM. A combination of family-based long-read whole-genome sequencing and short-read-based STR analysis identified the *TBC1D7* repeat expansion as a candidate locus in all three independent families described in this study. This finding reinforces the association between CCG-CGG expansions and a characteristic pattern of muscle weakness.

The clinical presentation of individuals with *TBC1D7* expansions was consistent with features reported in patients carrying expansions in other OPDM genes. However, we observed heterogeneity across families in terms of age at onset, initial symptoms, and disease severity. Family A presented with distal weakness in the fourth to fifth decade, whereas individuals in families B and C developed ptosis as the first symptom in the second decade. Family B showed a more aggressive disease progression, with early loss of independent ambulation, severe dysphagia and dysarthria, and respiratory involvement. As we did not observe a clear relationship between repeat size or sequence and disease severity, these findings suggest a role for additional modifiers and a possible interplay between genetic and epigenetic factors in shaping disease phenotype and course. Furthermore, these findings highlight how adopting broader inclusion criteria when screening for known OPDM genes or when searching for novel loci may increase diagnostic yield. Therefore, CCG-CGG expansions should be considered in individuals with partial OPDM phenotypes, including isolated ocular or distal weakness, particularly in the presence of supportive pathological features, such as rimmed vacuoles or p62-positive intranuclear inclusions on muscle biopsy.

Using large-scale control data, we delineated a pathogenic range of approximately 83–148 CCG repeat units for the *TBC1D7* expansion. Similar to other non-coding repeat expansion disorders, such as *RFC1*-associated CANVAS and *FGF14*-related SCA27B^24,25^, pathogenicity appears to depend on both repeat size and sequence composition. While most control alleles carry non-expanded or only minimally expanded CCGCTG motifs, the pathogenic CCG motifs in our study were located on a shared ancestral haplotype and appeared more prone to expansion. Although all previously reported OPDM-associated CCG expansions do not have a sequence change relative to the most common allele, our finding indicates that unbiased approaches are necessary when identifying novel tandem repeat loci.

Methylation as a disease-modifier has been reported for several OPDM-causing CCG repeat expansions and represents a well-established pathomechanism in *FMR1*^26^. The commonly accepted methylation threshold is 200 CCG units, although in the unaffected carrier C:I:1 we observe methylated reads as low as 125 CCG units, whereas in A:II:1 no methylation was detected, including four reads exceeding 200 CCG units. Similar observations have been reported in *FMR1,* where full-mutation carriers lacking methylation have been sporadically described^27^. In addition, pathogenic repeat lengths above the presumed methylation threshold have been reported for other OPDM-related loci, including *LRP12, NOTCH2NLC,* and *ABCD3.* These observations, although still anecdotal, suggest that the functional methylation threshold is not fixed but may be modulated by individual-specific factors. Somatic variability in repeat length or methylation status may represent one such modifier, although additional genetic or epigenetic influences are likely to contribute.

We further show that *TBC1D7* expansions are associated with increased transcript levels in patient-derived fibroblasts, whereas very large alleles are methylated and likely transcriptionally silenced. This pattern suggests, as for other OPDM-associated genes, that disease may be mediated by RNA and/or protein toxicity. Moreover, while homozygous loss-of-function variants in *TBC1D7* have been reported to cause a neurodevelopmental phenotype^31^, population data indicate that the gene is tolerant to haploinsufficiency.

Despite the heterogeneity of OPDM-associated loci, all currently identified pathogenic CCG repeat expansions appear to converge on a shared gain-of-function pathomechanism that is at least partly independent of the specific function of the repeat-containing gene. In *NOTCH2NLC*-related disease, for example, the expanded repeat resides within an upstream open reading frame and is translated into a polyglycine-containing protein that accumulates in p62-positive intranuclear inclusions and drives neurotoxicity in vivo^28^. Alternatively, or in addition, repeat-associated non-AUG (RAN) translation may contribute to protein toxicity, as recently shown for GGC repeats in *GIPC1*^29^. Polyglycine aggregates have been reported to induce mitochondrial dysfunction, disrupt nuclear lamina architecture, and alter RNA splicing, as demonstrated in GIPC1 iPSC-derived myotubes^29^. This convergence on a shared pathomechanism was recently demonstrated across different OPDM loci, including *GIPC1, RILPL1* and *LOC642361,* where the CCG-GGC repeat expansions were shown to be translated into polyglycine-containing proteins through upstream or alternative open reading frames^30^. Translation of these expanded repeats into polyglycine stretches stabilizes otherwise unstable micropeptides, resulting in their accumulation in p62-positive inclusions and inducing toxicity in muscle and neuronal models. Together, these findings further support a unifying gain-of-function mechanism across genetically heterogeneous OPDM-associated CCG repeat loci.

The identification of *TBC1D7* further strengthens emerging paradigms in the field of repeat expansion disorders. First, it supports a shift in the definition of pathogenicity in CCG-CGG repeat expansions from a strict threshold model to a pathogenic range model. In this framework, the lower boundary is defined by the minimum repeat size required to induce gain-of-function toxicity, which varies by locus, whereas the upper boundary is determined by methylation and gene inactivation.

Secondly, it reinforces a broader paradigm in Mendelian disorders associated with repeat expansions. In contrast to single-nucleotide variants, which are typically absent from control populations, pathogenic tandem repeats often arise at loci that are highly variable in the general population. Such variability may represent a prerequisite for further expansion and disease association, making highly polymorphic tandem repeats more likely to become pathogenic.

Thirdly, our findings suggest that pathogenic *TBC1D7* expansions occur on an ancestral, expansion-permissive haplotype that is associated with increased repeat variability at this locus. Such permissive haplotypes may predispose to repeat instability and facilitate pathogenic expansion. Consistent with this, the *TBC1D7* repeat appears to be germline-unstable, with both contraction (family C) and expansion (family A) observed following paternal transmission, thereby complicating genetic counselling for affected families. In conclusion, we identify a CCG expansion in *TBC1D7* as a novel cause of OPDM and related phenotypes. Our study highlights the power of combining family-based analyses with large patient and control datasets for the discovery of novel repeat expansions. It also underscores the utility of long-read sequencing for integrating repeat size, sequence composition, and methylation status in the analysis of CCG-CGG loci, emphasizing the need for systematic application of methylation-resolved long-read sequencing in unresolved neuromuscular disorders.

## Data availability statement

Participants’ sequencing data generated in this study cannot be made accessible due to the nature of the informed consent granting access to the researchers of this study only. The 1000 genomes ONT data is publicly accessible through https://registry.opendata.aws/1000-genomes. Sequencing data belonging to the 100,000 Genome Project used in this study is accessible and requests should be addressed to Genomics England Limited (https://www.genomicsengland.co.uk/research/academic/join-research-network). All other data generated is accessible upon reasonable request.

## Acknowledgements

We would like to acknowledge the help of Andy Wing Chun Pang at Bionano Genomics for his support in the analysis of the OGM data of A:II:1.

## References

1. Satoyoshi E, Kinoshita M. Oculopharyngodistal Myopathy Report of Four Families. JAMA Neurol. 1977;34;(2):89–92. doi:10.1001/archneur.1977.00500140043007.

2. Kumutpongpanich T, Ogasawara M, Ozaki A, et al. Clinicopathologic Features of Oculopharyngodistal Myopathy with LRP12 CGG Repeat Expansions Compared with Other Oculopharyngodistal Myopathy Subtypes. JAMA Neurol. 2021;78(7):853–863. doi:10.1001/jamaneurol.2021.1509

3. Ishiura H, Shibata S, Yoshimura J, et al. Noncoding CGG repeat expansions in neuronal intranuclear inclusion disease, oculopharyngodistal myopathy and an overlapping disease. Nat Genet. 2019;51(8):1222–1232. doi:10.1038/s41588-019-0458-z

4. Deng J, Yu J, Li P, et al. Expansion of GGC Repeat in GIPC1 Is Associated with Oculopharyngodistal Myopathy. Am J Hum Genet. 2020;106(6):793–804. doi:10.1016/j.ajhg.2020.04.011

5. Ogasawara M, Iida A, Kumutpongpanich T, et al. CGG expansion in NOTCH2NLC is associated with oculopharyngodistal myopathy with neurological manifestations. Acta Neuropathol Commun. 2020;8(1). doi:10.1186/s40478-020-01084-4

6. Zeng YH, Yang K, Du GQ, et al. GGC Repeat Expansion of RILPL1 is Associated with Oculopharyngodistal Myopathy. Ann Neurol. 2022;92(3):512–526. doi:10.1002/ana.26436

7. Cortese A, Beecroft SJ, Facchini S, et al. A CCG expansion in ABCD3 causes oculopharyngodistal myopathy in individuals of European ancestry. Nat Commun. 2024;15(1). doi:10.1038/s41467-024-49950-2

8. Gu X, Yu J, Jiao K, et al. Non-coding CGG repeat expansion in LOC642361/NUTM2B-AS1 is associated with a phenotype of oculopharyngodistal myopathy. J Med Genet. Published online 2023. doi:10.1136/jmg-2023-109345

9. Saito Y, Nishino I. Disease-specific genetic diagnostic strategies for muscle diseases unresolved by short-read sequencing. J Hum Genet. Springer Nature. Preprint posted online 2025. doi:10.1038/s10038-025-01391-5

10. 100,000 Genomes Pilot on Rare-Disease Diagnosis in Health Care — Preliminary Report. New England Journal of Medicine. 2021;385(20):1868–1880. doi:10.1056/NEJMoa2035790

11. Dolzhenko E, Bennett MF, Richmond PA, et al. ExpansionHunter Denovo: A computational method for locating known and novel repeat expansions in short-read sequencing data. Genome Biol. 2020;21(1):1–14. doi:10.1186/s13059-020-02017-z

12. Steyaert W, Sagath L, Demidov G, et al. Unraveling undiagnosed rare disease cases by HiFi long-read genome sequencing. Genome Res. 2025;35(4):755–768. doi:10.1101/gr.279414.124

13. van der Sanden B, Neveling K, Pang AWC, et al. Optical Genome Mapping for Applications in Repeat Expansion Disorders. Curr Protoc. 2024;4(7). doi:10.1002/cpz1.1094

14. De Coster W, Hoijer I, Bruggeman I, et al. Visualization and analysis of medically relevant tandem repeats in nanopore sequencing of control cohorts with pathSTR. Genome Res. Published online November 15, 2024:gr.279265.124. doi:10.1101/gr.279265.124

15. Isaac L, Xu GR, Pellerin D, Van De Vondel L, et al. The Genetic and Evolutionary Landscape of Pentanucleotide Tandem Repeats in Human. bioRxiv 2025.05.27.656002; doi: 10.1101/2025.05.27.656002

16. Auton A, Abecasis GR, Altshuler DM, et al. A global reference for human genetic variation. Nature. 2015;526(7571):68–74. doi:10.1038/nature15393

17. English AC, Dolzhenko E, Ziaei Jam H, et al. Analysis and benchmarking of small and large genomic variants across tandem repeats. Nat Biotechnol. Published online March 1, 2024. doi:10.1038/s41587-024-02225-z

18. Danzi MC, Xu IRL, Fazal S, et al. Detailed tandem repeat allele profiling in 1,027 long-read genomes reveals genome-wide patterns of pathogenicity. Preprint posted online January 7, 2025. doi:10.1101/2025.01.06.631535

19. Martin M, Patterson M, Garg S, et al. WhatsHap: fast and accurate read-based phasing. Preprint posted online November 2, 2016. doi:10.1101/085050

20. Yépez VA, Demidov G, Ellwanger K, et al. The Solve-RD Solvathons as a pan-European interdisciplinary collaboration to diagnose patients with rare disease. Nat Genet. 2025;57(10):2361–2370. doi:10.1038/s41588-025-02290-3

21. Dolzhenko E, English A, Dashnow H, et al. Characterization and visualization of tandem repeats at genome scale. Nat Biotechnol. Published online 2024. doi:10.1038/s41587-023-02057-3

22. Huang XR, Tang BS, Jin P, Guo JF. The Phenotypes and Mechanisms of NOTCH2NLC-Related GGC Repeat Expansion Disorders: a Comprehensive Review. Mol Neurobiol. Springer. 2022;59(1):523–534. doi:10.1007/s12035-021-02616-2

23. Fukuda H, Yamaguchi D, Nyquist K, et al. Father-to-offspring transmission of extremely long NOTCH2NLC repeat expansions with contractions: genetic and epigenetic profiling with long-read sequencing. Clin Epigenetics. 2021;13(1). doi:10.1186/s13148-021-01192-5

24. Cortese A, Simone R, Sullivan R, et al. Biallelic expansion of an intronic repeat in RFC1 is a common cause of late-onset ataxia. Nat Genet. 2019;51(4):649–658. doi:10.1038/s41588-019-0372-4

25. Pellerin D, Danzi MC, Wilke C, et al. Deep Intronic FGF14 GAA Repeat Expansion in Late-Onset Cerebellar Ataxia. New England Journal of Medicine. 2023;388(2):128–141. doi:10.1056/nejmoa2207406

26. Jacquemont S, Hagerman RJ, Hagerman PJ, Leehey MA. Review Fragile-X Syndrome and Fragile X-Associated Tremor/Ataxia Syndrome: Two Faces of FMR1. 2007. http://neurology.thelancet.comVol

27. Farmiloe G, Bejczy V, Tabolacci E, Willemsen R, Jacobs F. Transcriptomic profiling of unmethylated full mutation carriers implicates TET3 in FMR1 CGG repeat expansion methylation dynamics in fragile X syndrome. J Neurodev Disord. 2025;17(1). doi:10.1186/s11689-025-09609-5

28. Boivin M, Charlet-Berguerand N. Trinucleotide CGG Repeat Diseases: An Expanding Field of Polyglycine Proteins? Front Genet. Frontiers Media S.A. 2022;13. doi:10.3389/fgene.2022.843014

29. Jiao K, Chen X, Cao M, et al. Translation of GGC repeats into a toxic polyglycine protein 1 in oculopharyngodistal myopathy type 2 2. doi:10.1093/brain/awaf390/8295738

30. Boivin M, Yu J, Eura N, et al. GGC repeat expansions within new open reading frames are translated into toxic polyglycine proteins in oculopharyngodistal myopathy. Nat Genet. Published online February 17, 2026. doi:10.1038/s41588-026-02507-z

31. Alfaiz AA, Micale L, Mandriani B, et al. TBC1D7 Mutations are Associated with Intellectual Disability, Macrocrania, Patellar Dislocation, and Celiac Disease. Hum Mutat. 2014;35(4):447–451. doi:10.1002/humu.22529

